# Efficient maternofetal transplacental transfer of anti- SARS-CoV-2 spike antibodies after antenatal SARS-CoV-2 BNT162b2 mRNA vaccination

**DOI:** 10.1101/2021.03.11.21253352

**Authors:** Amihai Rottenstreich, Gila Zarbiv, Esther Oiknine-Djian, Roy Zigron, Dana G. Wolf, Shay Porat

**Affiliations:** Department of Obstetrics and Gynecology, Hadassah-Hebrew University Medical Center, Jerusalem, Israel; Clinical virology unit, Department of Clinical Microbiology and Infectious Diseases, Hadassah-Hebrew University Medical Center, Jerusalem, Israel

**Keywords:** pregnancy, vaccination, cord blood, serology, SARS-CoV-2

## Abstract

**Background:** Severe acute respiratory syndrome coronavirus 2 (SARS-CoV-2) during pregnancy and early infancy can result in severe disease. Evaluating the serologic response after maternal vaccination during pregnancy and subsequent transplacental antibody transfer has important implications for maternal care and vaccination strategies.

**Objective:** To assess maternal and neonatal SARS-CoV-2 antibody levels after antenatal mRNA vaccination.

**Design, Setting, and Participants:** This study took place at Hadassah Medical Center in Jerusalem, Israel in February 2021. Maternal and cord blood sera were collected for antibody measurement from mother/newborn dyads following antenatal vaccination.

**Exposure:** SARS-CoV-2 BNT162b2 mRNA vaccination.

**Main outcome and measures:** Spike protein (S) and receptor binding domain (RBD) - specific, IgG levels were evaluated in maternal and cord blood sera.

**Results:** The study cohort consisted of 20 parturients, with a median maternal age of 32 years and a median gestational age of 39^3/7^ weeks at the time of delivery. The median time lapsed from the first and second doses of vaccine administration until delivery was 33 [IQR 30-37] and 11 [IQR 9-15] days, respectively. Of the 20 dyads, all women an d infants were positive for anti S- and anti-RBD-specific IgG. Anti-S and anti-RBD-specific IgG levels in maternal sera were positively correlated to their respective concentrations in cord blood (ρ_s_= 0.72; P<0.001 and ρ_s_= 0.72; P <0.001, respectively). Anti-S and anti-RBD-specific IgG titers in cord blood were directly correlated with time lapsed since the administration of the first vaccine dose (ρ_s_= 0.71; P =0.001 and ρ_s_= 0.63; P=0.004, respectively).

**Conclusion and Relevance:** In this study, SARS-CoV-2 mRNA vaccine administered during pregnancy induced adequate maternal serologic response with subsequent efficient transplacental transfer. Our findings highlight that vaccination of pregnant women may provide maternal and neonatal protection from SARS-CoV-2 infection.

## Introduction

The rapidly emerging severe acute respiratory syndrome coronavirus 2 (SARS-CoV-2) has afflicted over 113 million individuals resulting in over 2.5 million deaths, since it was declared a pandemic by the World Health Organization on March 2020. The pressing need for effective tools to combat Coronavirus Disease 19 (COVID-19) has led to the accelerated development and recent approval of several targeted vaccines including two novel mRNA-based vaccines [1, 2]. A mass vaccination campaign using the BNT162b2 mRNA vaccine has commenced in Israel in December 2020.

Pregnant women are at higher risk for COVID-19 related illness [3, 4]. In addition, recent data show that severe SARS-CoV-2 infection is more common among infants as compared to older children [4, 5] Nevertheless, as pregnant women were excluded from the pivotal trials evaluating the aforementioned vaccines [1, 2], their safety and efficacy in the setting of pregnancy remain unknown.

Despite these uncertainties and given the risk for severe disease course, the Center for Disease Control and Prevention (CDC), the world Health Organization (WHO), and other agencies support offering pregnant women to receive the SARS-CoV-2 vaccine following shared decision making [6, 7]. Due to the paucity of literature and the high clinical relevance, we aimed to investigate the maternal serologic response after SARS-CoV-2 vaccination during pregnancy and its related subsequent transplacental antibody transfer.

## Methods

### Study Population

A prospective study following women admitted for delivery was performed in February 2021 at Hadassah Medical Center, a university affiliated hospital in Jerusalem, Israel. Women who received two doses of SARS-CoV-2 BNT162b2 mRNA vaccine during pregnancy were eligible for this study. Demographic and clinical data, were collected at the time of enrollment. The institutional review board of the Hadassah Medical Center approved this study (HMO-0064-21).

### Laboratory Methods

Following delivery, maternal and cord blood sera were collected for antibody measurement. Spike protein (S) (Liaison SARS-CoV-2 S1/S2 IgG, DiaSorin, Saluggia, Italy) and receptor binding domain (RBD)-specific (Architect SARS-CoV-2 IgG II Quant assay, Abbott Diagnostics, Chicago, USA), IgG levels were evaluated in maternal and cord blood sera. Maternal and cord blood sera were also tested for SARS-CoV-2 IgM (Liaison, DiaSorin, Saluggia, Italy).

### Statistical analysis

Patient characteristics are described as proportions for categorical variables and medians and interquartile range (IQR) for continuous variables without a normal distribution. Antibody levels and placental transfer ratios are expressed as medians and IQR. Correlations were reported using the Spearman’s test with the correspondent ρ_s_ and P values. The data were analyzed using Software Package for Statistics and Simulation (IBM SPSS version 24, IBM Corp, Armonk, NY).

## Results

During the study period, 20 parturients who received two doses of SARS-CoV-2 BNT162b2 mRNA vaccine were approached and agreed to participate. Median maternal age was 32 [IQR 28-37] years with a median gestational age of 39^3/7^ [IQR 38^2/7^-40^5/7^] weeks at the time of delivery. The median time lapsed from the first and second doses of vaccine administration until delivery was 33 [IQR 30-37] and 11 [IQR 9-15] days, respectively. Of the 20 dyads, all women and infants were positive for anti S- and anti-RBD-specific IgG. SARS-CoV-2 IgM antibodies were detected in 6 (30.0%) parturients, and were not detectable in any of the infants. Median SARS-CoV-2 anti-S and anti-RBD-specific IgG concentrations in maternal sera were 319 [IQR 211-1033] and 11150 [IQR 6154-17575] AU/mL, and 193 [IQR 111-260] and 3494 [IQR 1817-6163] AU/mL in cord blood, respectively.

The median placental transfer ratios of anti-S and anti-RBD specific IgG were 0.44 [IQR 0.25-0.61] and 0.34 [IQR 0.27-0.56], respectively.

SARS-CoV-2 anti-S and anti-RBD-specific IgG levels in maternal sera were positively correlated to their respective concentrations in cord blood (ρ_s_= 0.72; P<0.001 and ρ_s_= 0.72; P <0.001, respectively; Figure 1A, B). In addition, SARS-CoV-2 anti-S and anti-RBD-specific IgG titers in cord blood were directly correlated with increasing time since the first mRNA vaccine dose (ρ_s_= 0.71; P =0.001 and ρ_s_= 0.63; P=0.004, respectively; Figure 1C, D).

**Figure 1.**
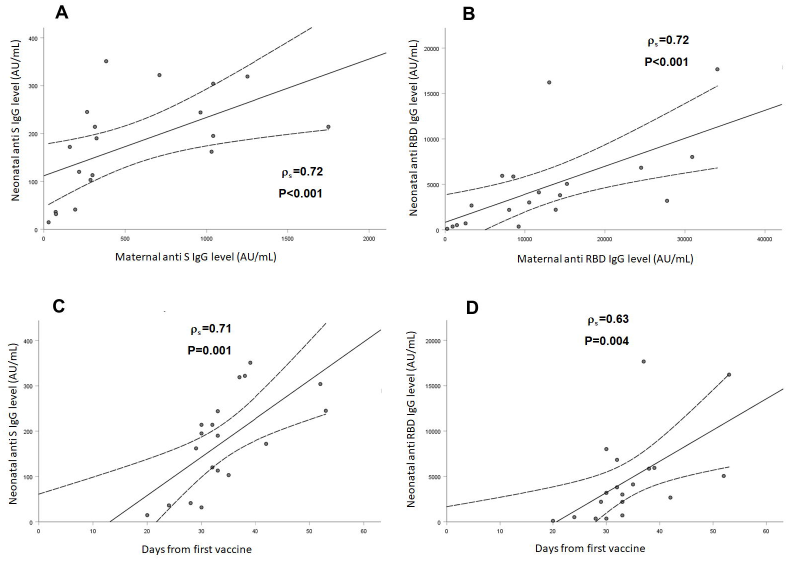
SARS-CoV-2 anti-S (A) and anti-RBD-specific (B) IgG levels in maternal sera were positively correlated to their respective concentrations in cord blood (ρ_s_= 0.72; P<0.001 and ρ_s_= 0.72; P <0.001, respectively). SARS-CoV-2 anti-S (C) and anti-RBD-specific (D) IgG titers in cord blood were directly correlated with increasing time since the first mRNA vaccine dose (ρ_s_= 0.71; P =0.001 and ρ_s_= 0.63; P=0.004, respectively). Correlations, as well as correspondent ρ_s_ and P values were calculated by Spearman’s test, as shown in each panel. The dotted lines are the 95% confidence intervals.

## Discussion

We measured SARS-CoV-2 anti-S and anti-RBD IgG in 20 mother/newborn dyads at a single center in Israel, following antenatal SARS-CoV-2 BNT162b2 mRNA vaccination. IgG antibodies were detected in all 20 maternal and cord blood sera. The current study finding may support the role of vaccination of pregnant women to induce both maternal and neonatal immunity.

Currently, there is lack of data regarding SARS-CoV-2 vaccination among pregnant women in terms of safety and efficacy. In addition, the degree of transplacental passive immunity induced by maternal SARS-CoV-2 vaccination is unestablished. In this regard, studies among pregnant women with SARS-CoV-2 infection reported conflicting results [8, 9], with some suggesting compromised transplacental transfer of naturally acquired antibodies [9], questioning the potential role of vaccination during pregnancy to confer neonatal protection against COVID-19.

Neonatal protection from infections is primarily dependent on maternally-derived, transplacentally acquired antibodies. We demonstrated an efficient placental transfer of IgG antibodies following maternal SARS-CoV-2 vaccination, and a positive correlation between maternal and cord blood antibody concentrations. Nevertheless, while neonatal antibody levels were satisfactory, placental transfer ratios were relatively lower as compared to prior studies of vaccine-elicited antibodies to influenza, pertussis, measles, rubella and hepatitis B, in which transfer ratios ranging from 0.8 to 1.7 have been reported [10, 11]. This concurs with a recent study among pregnant women with COVID-19 which also demonstrated diminished transplacental transfer of anti–SARS-CoV-2 IgG [9]. The mechanisms underlying this finding should be further investigated.

While the current study findings are promising, there are several questions which remain unanswered. First, the optimal timing for maternal vaccination is still unclear. In the current study, maternal and neonatal antibody levels were directly correlated to the time lapsed from vaccination, which is consistent with studies of respiratory syncytial virus vaccine given during pregnancy [12]. However, as all women in the current cohort were vaccinated during the third-trimester, whether earlier vaccination would result in similar antibody concentrations requires further evaluation. Based on the kinetics of the serologic response observed in pregnant women with COVID-19 and in non-pregnant subjects who received the SARS-CoV-2 mRNA vaccine, some authors have suggested that maternal vaccination during the early second trimester may the most ideal time to confer adequate maternal and neonatal immunity [12]. In addition, the durability of maternally-derived neonatal antibodies and the role of breastfeeding to further maintain neonatal immunity are other unsolved issues. Finally, larger studies are warranted to better assess the safety and efficacy of the different SARS-CoV-2 vaccines in the setting of pregnancy.

This study has several caveats, including its small sample size and single-site nature. In addition, the association between gestational age at delivery with transplacental transfer requires further investigation. Moreover, as previously stated, the effect of SARS-CoV-2 vaccination at different times throughout gestation remains to be explored.

Our findings demonstrate that antenatal SARS-CoV-2 vaccination induces an adequate maternal serologic response and has the potential to provide neonatal protection through transplacental transfer of vaccine-stimulated maternally-derived antibodies. These encouraging results have important implications for maternal care and the development of appropriate vaccination strategies. Further studies will be needed to better delineate the safety and efficacy of the different maternal SARS-CoV-2 vaccines available and better define transplacental antibody dynamics at earlier gestational ages.

## Data Availability

The data that support the findings of this study are available from the corresponding authors, upon reasonable request.

## Authors’ contributions

Dr Rottenstreich had full access to all of the data in the study and takes responsibility for the integrity of the data and the accuracy of the data analysis.

Concept and design: Porat, Rottenstreich, Wolf.

Acquisition, analysis, or interpretation of data: All authors.

Drafting of the manuscript: Rottenstreich, Zigron, Zarbiv, Porat, Wolf.

Laboratory analyses: Oiknine-Djian, Wolf.

Statistical analysis: Rottenstreich.

All authors read and approved the final manuscript.

## Additional Contributions

We thank Dr. Doron Kabiri, Dr. Shlomi Yahalomi, Prof. Yosef Ezra, Dr. Gali Gordon, Dr. Naama Lessens and Dr. Roy Alter for their assistance in patients’ enrollment. We also thank Rimma Barsuk and Yulia Yachnin for their technical assistance..

## References

1. Polack FP, Thomas SJ, Kitchin N, et al. Safety and efficacy of the BNT162b2 mRNA Covid-19 vaccine. N Engl J Med 2020;383:2603–2615.

2. Baden LR, El Sahly HM, Essink B, et al. Efficacy and safety of the mRNA-1273 SARSCoV-2 vaccine. N Engl J Med 2021; 384:403–416.

3. Zambrano LD, Ellington S, Strid P, et al. Update: characteristics of symptomatic women of reproductive age with laboratory-confirmed SARS-CoV-2 infection by pregnancy status - United States, January 22-October 3, 2020. MMWR Morb Mortal Wkly Rep 2020;69:16 41– 7.

4. Woodworth KR, Olsen EO, Neelam V, et al. CDC COVID-19 Response Pregnancy and Infant Linked Outcomes Team; COVID-19 Pregnancy and Infant Linked Outcomes Team (PILOT). Birth and infant outcomes following laboratory-confirmed SARS-CoV-2 infection in pregnancy: SET-NET, 16 jurisdictions, March 29-October 14, 2020. MMWR Morb Mortal Wkly Rep. 2020;69(44):1635–1640.

5. Kim L, Whitaker M, O’Halloran A, et al; COVID-NET Surveillance Team. Hospitalization rates and characteristics of children aged <18 years hospitalized with laboratory-confirmed COVID-19—COVID-NET, 14 states, March 1-July 25, 2020. MMWR Morb Mortal Wkly Rep. 2020;69(32):1081 –1088.

6. Centers for Disease Control and Prevention. COVID-19 (coronavirus disease): people with certain medical conditions. Available at: https://www.cdc.gov/coronavirus/2019-ncov/need-extra-precautions/people-with-medical-conditions.html. Accessed February 24, 2021.

7. Stafford IA, Parchem JG, Sibai BM. The coronavirus disease 2019 vaccine in pregnancy: risks, benefits, and recommendations. Am J Obstet Gynecol 2021; 30;S0002-9378(21)00077-6.

8. Flannery DD, Gouma S, Dhudasia MB, et al. Assessment of maternal and neonatal cord blood SARS-CoV-2 antibodies and placental transfer ratios. JAMA Pediatr 2021;

9. Edlow AG, Li JZ, Collier AY, et al. Assessment of maternal and neonatal SARS-CoV-2 viral load, transplacental antibody transfer, and placental pathology in pregnancies during the COVID-19 pandemic. JAMA Netw Open. 2020;3(12):e2030455.

10. Post AL, Li SH, Berry M, et al. Efficiency of placental transfer of vaccine-elicited antibodies relative to prenatal Tdap vaccination status. Vaccine. 2020;38(31):4869–4876.

11. Munoz FM, Patel SM, Jackson LA, et al. Safety and immunogenicity of three seasonal inactivated influenza vaccines among pregnant women and antibody persistence in their infants. Vaccine. 2020;38(33):5355–5363.

12. Munoz FM. Can We Protect Pregnant Women and Young Infants From COVID-19 Through Maternal Immunization? JAMA Pediatr. 2021;

